# Detection of anti-nucleocapsid antibodies associated with primary SARS-CoV-2 infection in unvaccinated and vaccinated blood donors

**DOI:** 10.1101/2024.05.23.24307822

**Authors:** Eduard Grebe, Mars Stone, Bryan R. Spencer, Akintunde Akinseye, David Wright, Clara Di Germanio, Roberta Bruhn, Karla G. Zurita, Paul Contestable, Valerie Green, Marion C. Lanteri, Paula Saa, Brad J. Biggerstaff, Melissa M. Coughlin, Steve Kleinman, Brian Custer, Jefferson M. Jones, Michael P. Busch

**Author notes:** These authors contributed equally to this work (Eduard Grebe, Mars Stone).

## Abstract

Anti-nucleocapsid (N) antibody assays can be used to estimate SARS-CoV-2 infection prevalence in regions employing anti-spike based COVID-19 vaccines. However, poor sensitivity of anti-N assays in detecting infections after vaccination (VI) has been reported. To support serological monitoring of infections, including VI, in a large blood donor cohort (N=142,599), we derived a lower cutoff for identifying previous infection using the Ortho VITROS Anti-SARS-CoV-2 Total-N Antibody assay, improving sensitivity while maintaining specificity >98%. Sensitivity was validated in samples donated after self-reported infections diagnosed by a swab-based test. Sensitivity for first infections in unvaccinated donors was 98.1% (95% CI: 98.0,98.2) and for VI was 95.6% (95.6,95.7), using the standard cutoff. Regression analysis showed sensitivity was reduced in the Delta compared to Omicron period, in older donors, asymptomatic infections, ≤30 days after infection and for VI. The standard Ortho anti-N threshold demonstrated good sensitivity, which was modestly improved with the revised cutoff.

## Introduction

In the United States (U.S.), like many countries, convenience sample serosurveillance studies (e.g., in blood donors) have demonstrated that a large majority of the population have antibodies to SARS-CoV-2 as a result of vaccination, infection, or both (1–3). Therefore, continued surveillance using serology requires robust detection of first infections that occur in vaccinated individuals (vaccination followed by infection, VI) and reinfections to yield meaningful estimates of infection incidence. Serological detection of anti-nucleocapsid (anti-N) antibodies (Abs) has been a critical tool for detection of previous SARS-CoV-2 infection and to discriminate between vaccine- and infection-induced Ab reactivity in the context of spike (S)-based vaccines.

The National Blood Donor Cohort (NBDC), a longitudinal study sponsored by the U.S. Centers for Disease Control and Prevention (CDC), was conducted in partnership with the two largest U.S. blood collectors, Vitalant and the American Red Cross (ARC), and their central testing laboratory Creative Testing Solutions (CTS) (4). Participating donors were classified into four groups based on infection and vaccination status as of mid-2021: Not previously infected or vaccinated, previously infected, previously vaccinated, or both previously infected and vaccinated. An earlier iteration of this program (the National Blood Donor Serosurvey, NBDS) executed serial monthly cross-sectional serosurveys from July 2020 through December 2021 (5–7), to provide population-weighted seroprevalence estimates. However, as vaccination rates increased in 2021, the proportion of donations with vaccine- and/or infection-induced anti-S reactivity approached 95%, and rates of infection-induced anti-N reactivity exceeded 75% in the U.S. by the end of 2022 (1, 8), thus diminishing the value of cross-sectional serosurveillance. Important objectives of the NBDC included continued monitoring of SARS-CoV-2 infection incidence and vaccine- and infection-induced seroprevalence, in the context of endemic SARS-CoV-2 transmission and increasing frequency of VI and reinfections (Stone et al., forthcoming).

Several studies have suggested that the sensitivity of anti-N serology for detecting previous SARS-CoV-2 infection is reduced in vaccinated relative to unvaccinated people (9–12). Significantly reduced anti-N Ab reactivity has been reported in previously vaccinated individuals with PCR-confirmed infections, compared to infections in unvaccinated individuals (9). Moderna mRNA vaccine trial data showed lower rates of anti-N seropositivity following PCR-confirmed infection among vaccine compared to placebo recipients (40.4% vs 93.4%) (10). A blunted anti-N Ab response following VI, and consequently reduced sensitivity of anti-N serology, may result from suppression of viral replication due to existing anti-S Abs and an associated anamnestic response (13). Additionally, studies relying on anti-N IgG Ab detection (14) may be confounded by rapidly waning Abs below the limit of detection (seroreversion) (15–18).

We previously demonstrated good performance of the Ortho VITROS Immunodiagnostic Products Anti-SARS-CoV-2 Total N Antibody (Ortho anti-N total Ig) and Roche Elecsys NC anti-SARS-CoV-2 (Roche anti-N total Ig) assays for serosurveillance applications, without differentiating infections in vaccinated and unvaccinated people (19). In the present study, to increase sensitivity while maintaining high specificity for serological detection of VI, we derived a revised reactive/non-reactive cutoff for these assays. In addition, for the Ortho anti-N total Ig assay we sought to validate the sensitivity of both the manufacturer’s recommended and our revised cutoff for identifying first infections in vaccinated and unvaccinated donors who self-reported swab-confirmed infections (SCIs). We then assessed factors influencing detection of anti-N Abs and assessed the durability of Ab detection following primary infection.

## Materials and Methods

### Study population

Repeat blood donors were identified from two national blood collection organizations (Vitalant and ARC) who had known prior SARS-CoV-2 infection and COVID-19 vaccination status determined during June 2020-July 2021, when all donations were tested for anti-SARS-CoV-2 Ab and donors reported vaccination status at the time of donation. Eligible donors were those presenting at least twice during the screening period and meeting all blood donor eligibility criteria. The NBDC includes 142,599 repeat blood donors. Eligibility screening was based on donations tested with Ortho VITROS anti-SARS-CoV-2 (Ortho anti-S total Ig) and Roche anti-N total Ig assays, and all anti-S reactive samples were retained (6, 7, 20, 21). During follow-up from July 2021 to December 2022, donation specimens were identified in real time and stored frozen at -20°C. During 2022, one donation specimen per donor per quarter was tested using the Ortho VITROS Anti-SARS-CoV-2 IgG Quantitative test (Ortho anti-S IgG) and the Ortho anti-N total Ig assay at CTS and Vitalant Research Institute. Self-reported vaccination status was captured at each donation as part of routine donation procedures. All cohort donors were invited to respond to electronic surveys on vaccination and infection history, and clinical outcomes of infections, with an overall response rate of 46.5%. NBDC seroprevalence estimates have been published (1).

### Analysis and statistical methods

All analyses were conducted using the SAS System, Version 9.4 (SAS Institute Inc., Cary, NC, USA). Specific statistical analyses are described in each subsection below.

#### Derivation of a revised non-reactive/reactive cutoff for the Ortho anti-N assay

To detect infections serologically with optimal sensitivity, we derived a revised cutoff using receiver operating characteristic (ROC) curve analysis (**Supplementary Appendix**), with gray zone reactivity consequently defined as reactivity above the revised threshold and below the standard threshold (0.395≤signal-to-cutoff ratio [S/CO]<1.0).

#### Impact of vaccination status on anti-N reactivity

To assess whether first infections that were VI were associated with reduced post-infection anti-N reactivity, compared to first infections in unvaccinated donors, we evaluated Ortho anti-N assay reactivity distributions following infection in two groups: (1) serologically-identified putative first infections, defined as first donation sample for each donor in which anti-N reactivity was above the revised cutoff, amongst all donors in the NBDC, and (2) first survey-reported swab-confirmed infections (SCIs). In (1), vaccination status was based on self-report at the time of donation. In (2), SCIs were defined as infections confirmed by viral antigen or PCR testing or by physician diagnosis (presumed positive swab-based test) and vaccination status at the time of infection was based on survey-reported vaccinations. In both groups, donation samples were stratified by vaccination status (vaccinated/unvaccinated) at the time of infection and by time period: the Delta variant era, July–December 2021, and the Omicron era, January– December 2022 (22), and further stratified by quarter. The proportions of donation samples with reactivity in the gray zone were calculated each group.

#### Sensitivity of manufacturer’s recommended and revised cutoffs for detection of first infections

To validate sensitivity, we identified survey-reported swab-confirmed first SARS-CoV-2 infections, defined as above. Infections were classified as occurring in an unvaccinated donor if the donor had not reported any vaccination prior to the date of diagnosed infection; and as a VI if it occurred ≥14 days after completion of an approved primary vaccination series (one dose of the Janssen vaccine or two doses of either the Pfizer-BioNtech or Moderna mRNA vaccines). For cases to be included in this analysis, ≥1 donation sample had to have been collected 14-180 days post diagnosis with no prior anti-N reactivity above the standard cutoff. A total of 2,751 swab-confirmed first infections in unvaccinated donors and 8,187 swab-confirmed first infections that were VIs were identified. For a secondary, more restrictive, analysis, VIs were only included if a post-vaccination anti-S reactive, anti-N non-reactive sample had been collected before infection, demonstrating vaccine-induced anti-S Ab seroconversion in the absence of infection-induced anti-N Abs. Infections occurring after only one mRNA vaccination dose or <14 days after completion of a vaccination series were excluded. For this analysis, 5,079 VI cases were identified. Only one post-infection sample per case was included in either analysis.

We estimated the sensitivity of both the manufacturer’s recommended and our revised cutoffs on the Ortho anti-N assay in first donation samples following SCI, stratified by vaccination status of the donor at the time of infection. Additionally, we stratified infections according to the variant era (Delta period *vs.* Omicron period), donor age (<65 years *vs.* ≥65 years), and whether the infection was associated with ≥1 self-reported symptom. Sensitivity was defined as the proportion of samples that were reactive, with the 95% confidence interval (CI) calculated using the Wilson score method. Differences in sensitivity for different strata were assessed using the binomial exact test. Differences in sensitivity associated with different cutoffs computed on the same stratum were assessed by computing a p-value for the difference in the Youden’s Js associated with each cutoff.

#### Factors associated with anti-N seroconversion following swab-confirmed infection

Bivariate and multivariable logistic regression was performed to assess the impact of vaccination status, timing of sample collection relative to infection, donor demographics (age and gender), presence of symptoms and variant era on anti-N Ab detection. Samples collected <14 days or >180 days after SCI were included because the model adjusted for time from infection to sample collection. Unadjusted and adjusted odds ratios (ORs) from logistic regression were computed. After assessing unadjusted ORs, the vaccination status and timing variables were combined for the multivariable regression.

#### Durability of anti-N Ab detection

Durability of anti-N detection after primary infection was assessed in unvaccinated and vaccinated donors by examining the proportion of primary infections detectable by time from SCI to sample collection (0-13, 14-30, 31-60, 61-90, 91-180, 181-365 and >365 days). To account for multiple observations per time bin, observations were weighted so that donors were equally weighted within each time bin, regardless of the number of observations. Confidence intervals were computed using the Wilson score method.

#### Impact of adjustment for anti-N sensitivity on seroprevalence estimates

To assess the potential impact of imperfect sensitivity and specificity on estimates of infection rates among vaccinated individuals, we compared adjusted and unadjusted estimates of the proportion of vaccinated donors (not previously infected) who experienced VI during three time periods in the NBDC: Q2 2021-Q1 2022, Q1-Q2 2022 and Q2-Q3 2022. As we are not able to estimate the proportion of infections that were asymptomatic from survey data since diagnostic testing is largely driven by the presence of symptoms, for the purposes of this model we used an estimated proportion of VIs that are asymptomatic of 32.4% (23). We then adjusted for a weighted average of symptomatic- and asymptomatic-specific sensitivity estimates, as well as for specificity estimated using prepandemic samples. The adjusted proportion of individuals infected during each time period was computed using the estimator derived by Rogan and Gladen (24), and 95% confidence intervals were based on parametric bootstrapping (10,000 iterations) of the proportion of tests that were reactive, sensitivity and specificity (treated as binomially distributed and incorporating the uncertainty arising from limited sample size).

## Results

### Revised cutoff for detecting previous infection using Ortho anti-N assay

The non-reactive/reactive threshold on the Roche anti-N cutoff index (COI) that maximized Youden’s J was COI≥0.205, and this optimized cutoff was used in defining cases for the Ortho ROC curve analysis. The ROC-optimized threshold on the Ortho anti-N assay was S/CO≥0.395, which had a sensitivity of 98.7% and a specificity of 98.7% in the Ortho optimization sample set. The area under the ROC curve was 0.994. See **Supplementary Appendix** for additional detail.

### Impact of vaccination status on anti-N reactivity

Distributions of Ortho anti-N assay S/COs in first longitudinal samples with reactivity above the revised cutoff (S/CO ≥0.395, i.e., putative first infections) from previously uninfected donors (based on negative Ortho anti-N results (S/CO<0.395) in all previous longitudinal samples), by vaccination status and variant era are shown in **Figure 1**. Serologically identified first infections in 6,555 unvaccinated donors and in 22,217 vaccinated donors are shown in the left and right panels respectively of **Figure 1A**. **Figure 1B** shows the Ortho anti-N assay S/CO distributions following self-reported first SCIs in previously uninfected donors (as indicated by self-report and anti-N serology), by vaccination status and variant era. A total of 2,751 SCIs in unvaccinated donors and 8,187 swab-confirmed VIs are shown in the left and right panels respectively.

**Figure 1.**
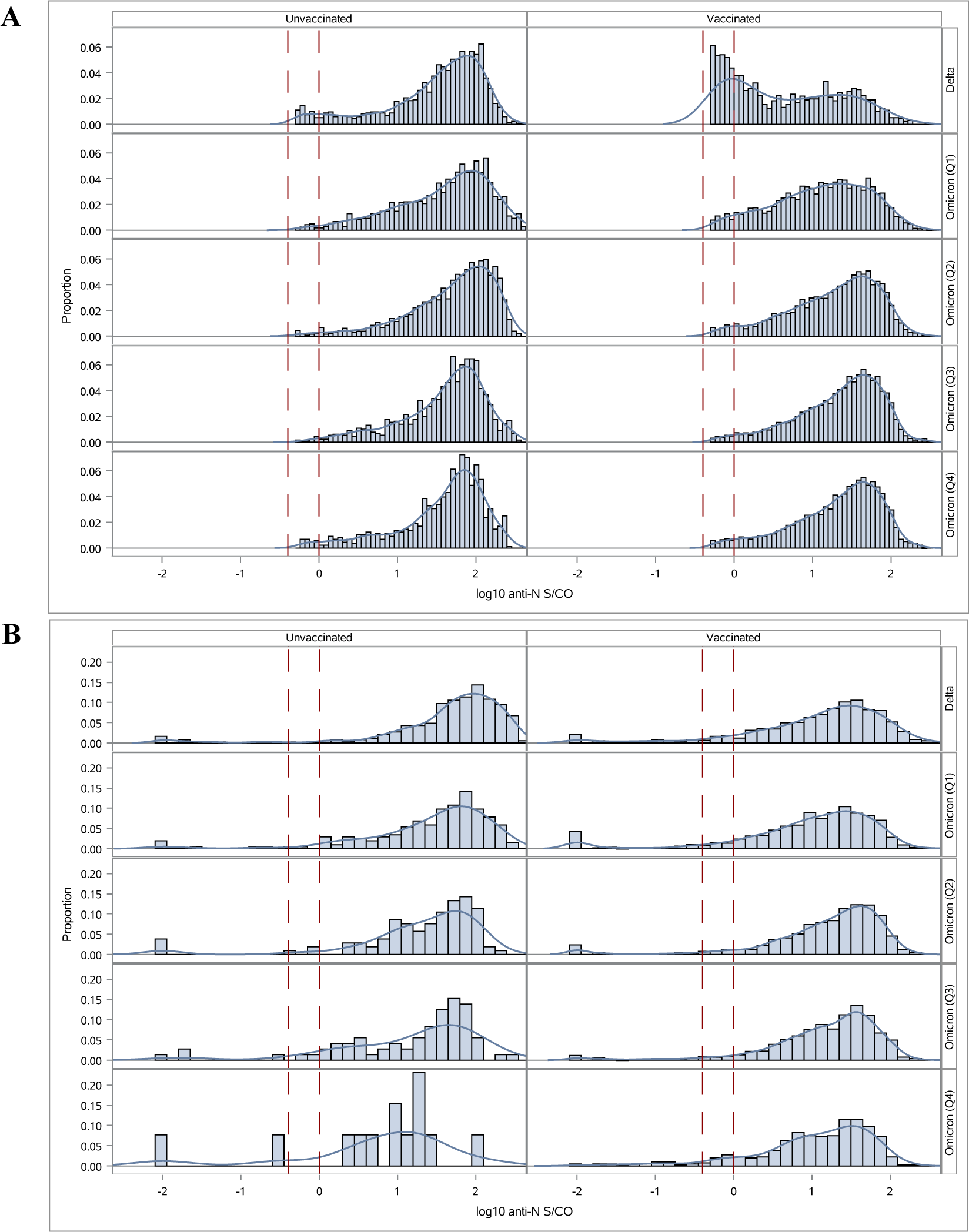
Ortho anti-nucleocapsid (anti-N) signal intensity distributions observed in vaccinated and unvaccinated donors after primary SARS-CoV-2 infection. Panel A shows reactivity of putative serologically identified infections at the first longitudinal sample showing reactivity above the reduced cutoff (gray zone reactivity, S/CO≥0.395≤1), by vaccination status and variant era (6,555 unvaccinated donors and 22,217 vaccinated donors). Panel B shows reactivity at the first sample collected after reported swab-confirmed infection (14 to 180 days post-infection), by vaccination status and variant era (2,751 unvaccinated donors and 8,187 vaccinated donors). Vertical dashed lines indicate the gray zone of anti-N.

During the Delta era, 35.2% of serologically identified VIs showed gray zone reactivity (0.395≤S/CO<1.0) compared to 7.5% of serologically identified primary infections in unvaccinated donors, declining to 3.8% and 2.7% respectively by the fourth quarter of 2022 (Omicron period Q4, **Figure 1A)**. Among survey respondents with SCI there was not a similar increased proportion with gray zone anti-N reactivity in the Delta period, with the vast majority having reactivity above the standard cutoff **(Figure 1B)**.

### Sensitivity for detection of swab-confirmed primary infections

Overall sensitivity of the manufacturer’s cutoff for the Ortho anti-N assay was 98.1% (95% confidence interval [CI]: 98.0–98.2) for detection of first infections in unvaccinated donors and 95.6% (95.6–95.7) for detection of first infections that were VI (**Table 1)**. Sensitivity was increased when using the revised cutoff, to 98.4% (98.4–98.5) in unvaccinated donors and to 97.0% (96.9–97.0) for detection in VI. While sensitivity is necessarily increased by reducing the cut-off from the manufacturer’s suggestion, Youden’s J index is not statistically improved by the reduced cut-off (p=0.13 using a single-tailed test). Sensitivity for detection of VI using the standard cutoff was higher during the Omicron era (96.0%, 95.9–96.0) than the Delta era (93.9%, 93.5–95.4), p=0.001; and was higher for detecting symptomatic VI than asymptomatic VI – 96.2% (96.1–96.2) *vs.* 90.1% (88.5–91.7), p<0.0001. A secondary sensitivity analysis using a more restrictive case definition of VI, that required anti-S seroconversion following vaccination, showed similar patterns (**Supplementary Table 1**).

**Table 1.**
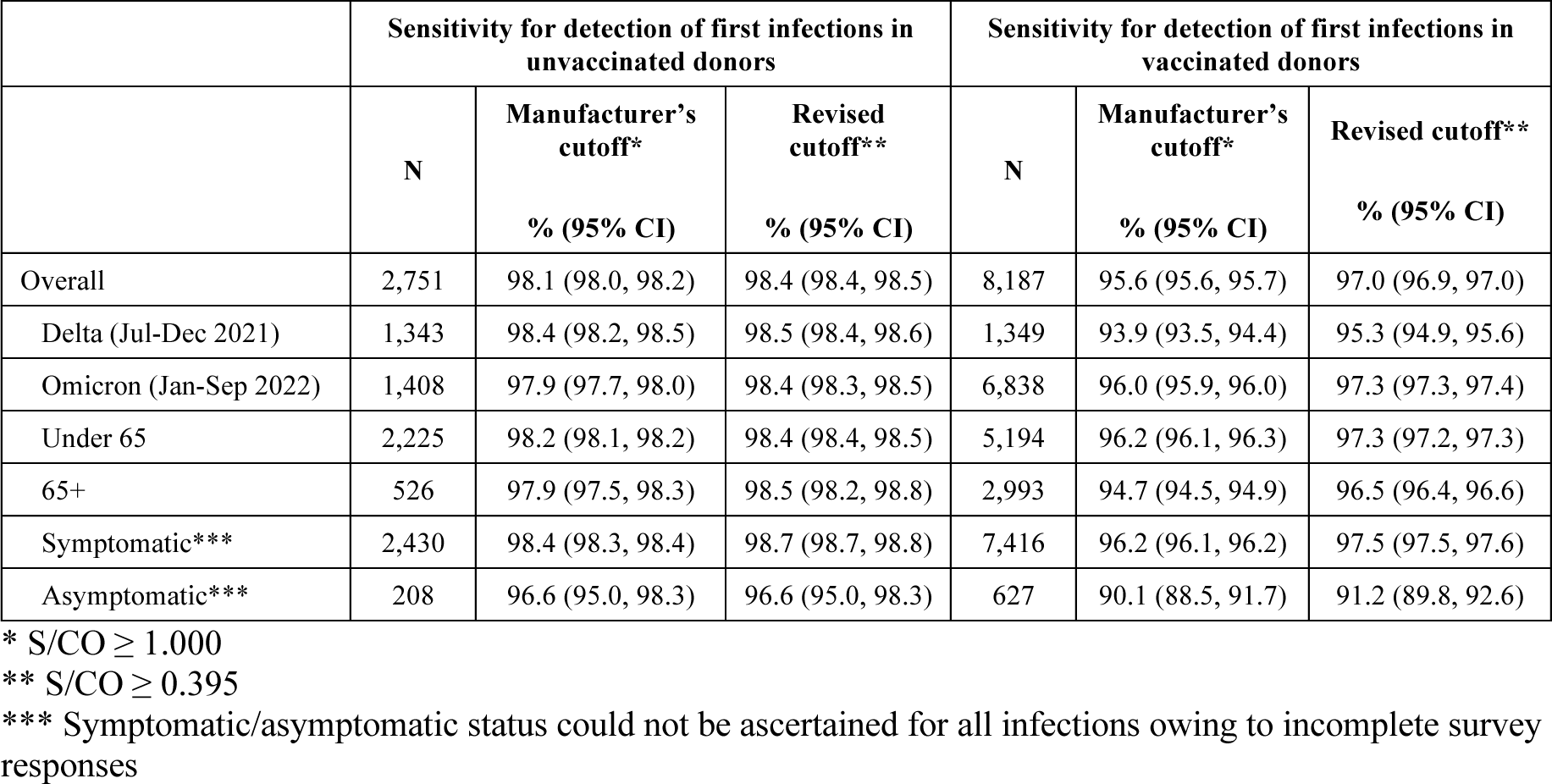
Sensitivity of Ortho anti-nucleocapsid assay for detection of first infections in unvaccinated and vaccinated donors. Proportion reactive in the first sample collected after reported swab-confirmed infection, collected 14 to 180 days post-infection.

### Factors associated with anti-N seroconversion following swab-confirmed infection

Using first samples collected after first SCIs, bivariate logistic regression showed that infection during the Delta era, age <65 years, female gender, symptomatic infection, being unvaccinated at the time of infection, longer time between vaccination and infection, and longer intervals between infection and sample collection, were all statistically significantly associated with increased probability of detection (see **Figure 2** and **Supplementary Table 2**). Notably, the percent detected in all time to sample categories, apart from donation samples collected <14 days post-infection (48.9%), ranged from 87.7% to 98.1%.

**Figure 2.**
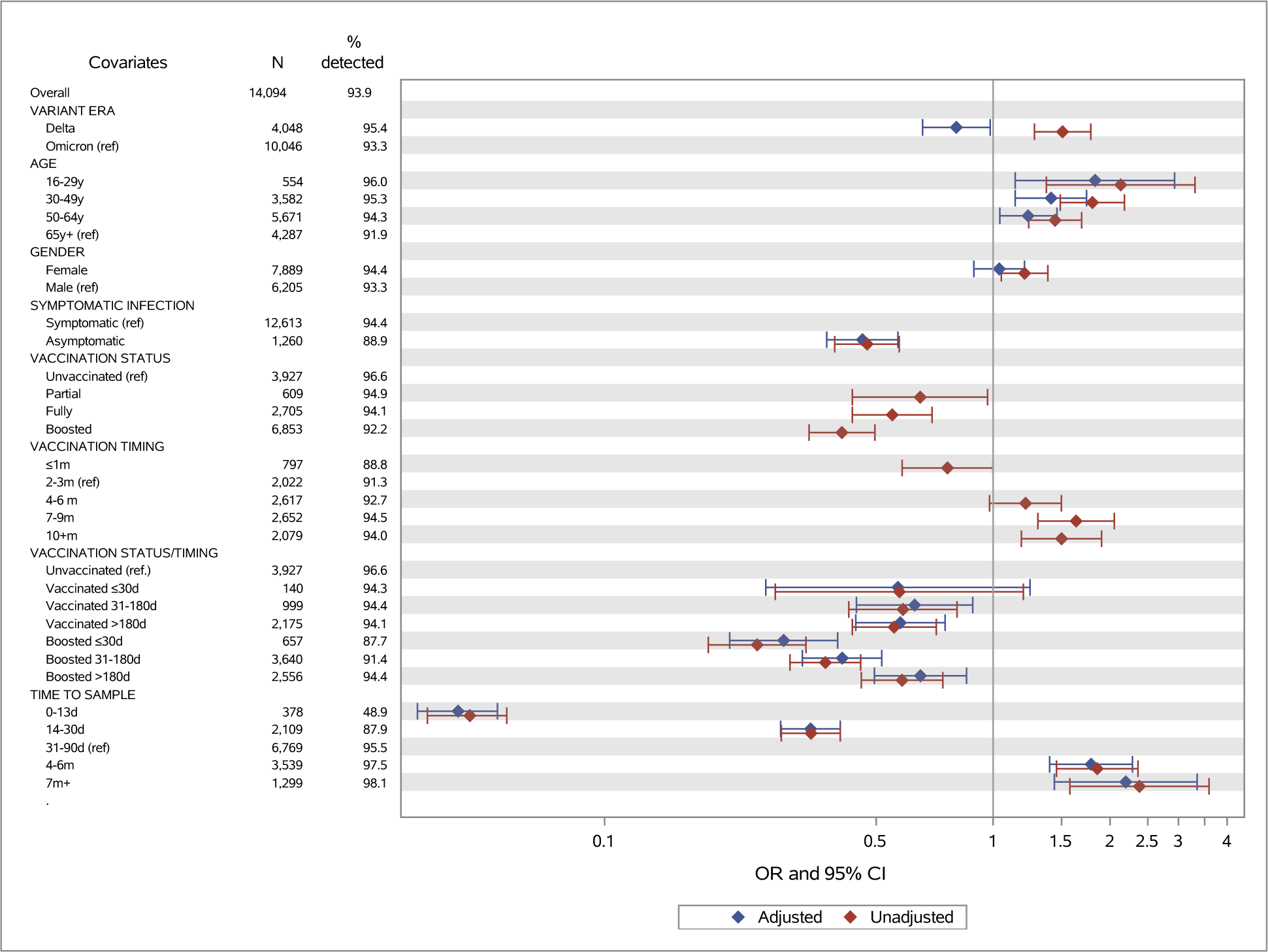
Factors influencing anti-nucleocapsid (anti-N) seroconversion following swab-confirmed first SARS-CoV-2 infections. The number of samples in each group and the proportion of anti-N reactive are shown in the table and odds ratios (ORs) and 95% confidence intervals (CI) from logistic regression are shown graphically, with unadjusted ORs in red and adjusted ORs in blue. In the multivariable regression model (adjusted ORs), the categories for certain variables have been grouped together: the vaccination status at the time of infection and timing of most recent vaccine prior to infection were combined in the variable VACCINATION STATUS/TIMING, and in the variable for time from infection to tested sample, the groups for samples collected 7-12 months and >1 year post infection were combined. ORs are also shown in Supplementary Table 2.

In multivariable logistic regression, infection during the Delta period (*vs.* Omicron period) remained statistically significant, but the direction of impact changed to reduced detection (adjusted odds ratio [aOR]=0.80, 95% CI 0.66–0.98) from increased detection in bivariate analysis (odds ratio [OR]=1.51, 95% CI 1.28–1.78), possibly because variant era (calendar time) is also strongly associated with vaccination status. Younger age groups had higher odds of detection than donors aged 65 years and older, while donor gender was not significantly associated with detection in the multivariable analysis. Asymptomatic infection was significantly associated with reduced detection (aOR=0.46, 95% CI 0.37–0.57). Being vaccinated at the time of infection significantly reduced detection compared to being unvaccinated (aORs<1), with the exception of primary vaccination ≤30 days before infection, which was not statistically significant. Vaccination reduced detection compared to no vaccination, and more recent receipt of either a primary vaccination series (31-180 days before infection) or a booster vaccination (≤30 days or 31-180 days before infection) was associated with reduced odds of detection than when infections occurred more than 180 days since the last vaccine. Compared to sample collection 31-90 days post-infection, sample collection less than fourteen days and 14-30 days after infection were associated with greatly reduced detection (aOR=0.04, 95% CI 0.03–0.05 and 0.34, 95% CI 0.28–0.40, respectively), while sample collection 3-6 months or 7 or more months after infection were associated with increased detection (aOR=1.79, 95% CI 1.40–2.28 and 2.19, 1.44–3.35). Percentage detected in each category, and unadjusted and adjusted ORs are shown in **Figure 2** and provided in **Supplementary Table 2**.

### Durability of anti-N Ab detection

Anti-N reactivity was detected in less than half of specimens collected <14 days post-infection, more than 80% of specimens collected 14-30 days post-infection and more than 90% of specimens collected >90 days post-infection, in both donors who were vaccinated and unvaccinated at the time of infection. No decline in percent detected was observed in later time bins, including >1 year post-infection days. The proportion detected was slightly lower in vaccinated donors, and for asymptomatic infections at all times post-infection (**Figure 3)**.

**Figure 3.**
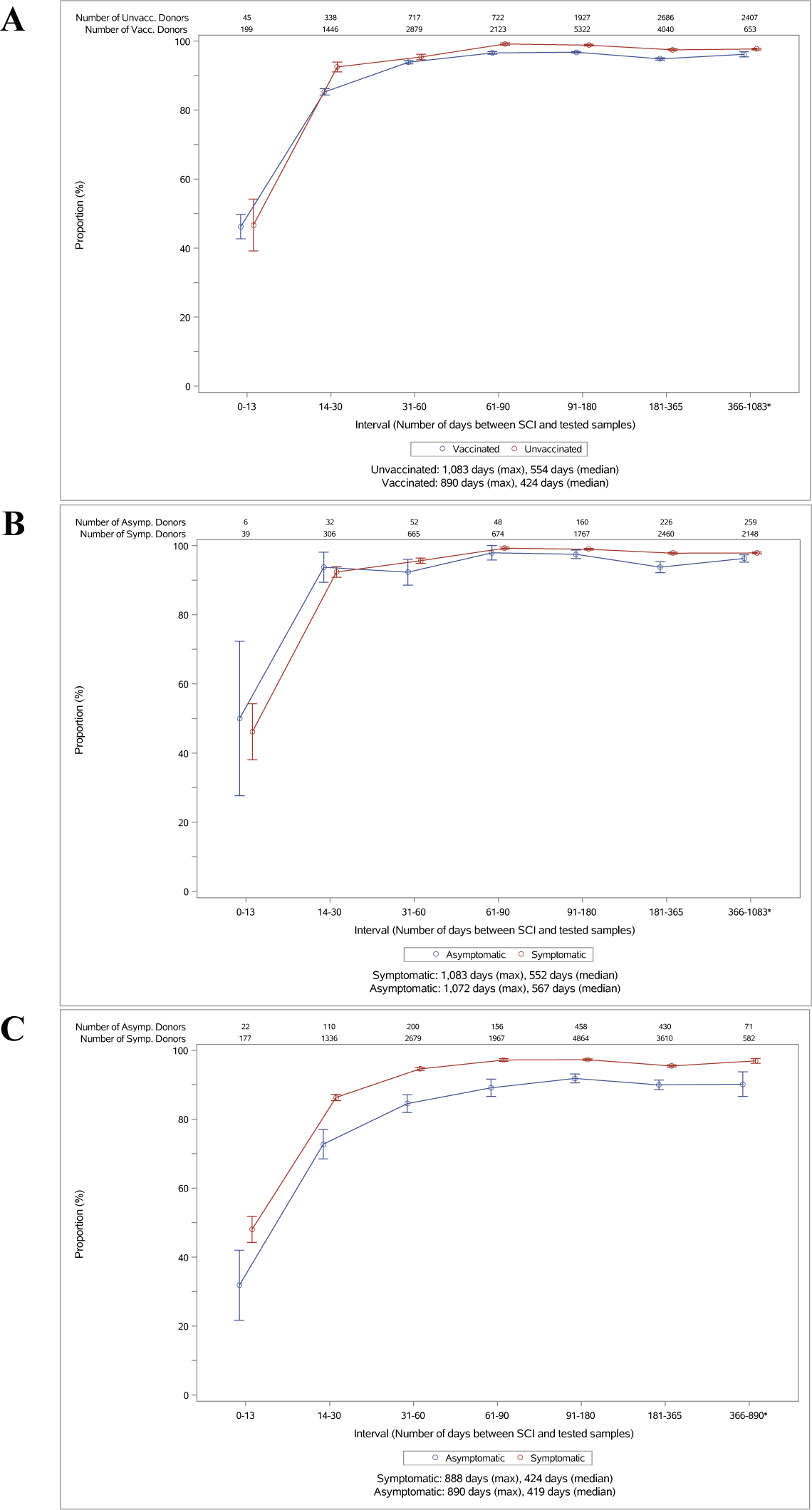
Sensitivity of anti-nucleocapsid serology by time from swab-confirmed infection (SCI) to sample collection in vaccinated and unvaccinated donors, using the manufacturer’s recommended cutoff. The figure shows proportion of donors showing reactivity in first or subsequent samples following SCI. To account for multiple observations per time bin, observations were weighted so that donors were equally weighted within each time bin, regardless of the number of observations. Panel A shows reactive proportions stratified by vaccination status at the time of infection. Panel B is restricted to infections in unvaccinated donors stratified by reported symptoms, and Panel C shows reactive proportions in unvaccinated donors with SCIs stratified by reported symptoms. The medium and maximum durations of follow-up for each group are provided at the bottom of each panel.

### Impact of adjustment for anti-N sensitivity on seroprevalence estimates

Estimates of the proportion of vaccinated donors who had become infected in each time period (using the anti-N test), adjusted for sensitivity and specificity, and assuming that 32.4% of infections were asymptomatic, differed little from unadjusted estimates. Adjusted estimated infection rates increased in each time period by 0.2 to 0.9 percentage points, or proportionally by 1.5-4% (**Supplementary Table 3**).

## Discussion

Despite reports of sensitivity as low as 40% for serological detection of VI (10), our study demonstrates sensitivity for detection of swab-confirmed first infections >98% and >95% for unvaccinated and vaccinated individuals respectively. Additionally, this study supports use of the manufacturer’s recommended cutoff for identifying previous infections in both vaccinated and unvaccinated individuals. Timing of sample collection after infection impacted sensitivity, with poorer sensitivity <30 days post-infection, thus timing of sample collection must be considered when interpreting previous reports.

In our validation of sensitivity for detection of first infections, the number of infections in vaccinated donors greatly exceeded that in unvaccinated donors in the study period, a function of high donor vaccination rates. This demonstrates the importance of sensitive detection of VI in SARS-CoV-2 serosurveillance programs.

The revised cutoff for the Ortho anti-N assay offered minimal improvement in sensitivity to detect VI compared to the manufacturer’s cutoff. Our ROC analysis equally weighted sensitivity and specificity, an approach appropriate for surveillance applications, but potentially less appropriate for clinical applications prioritizing specificity. The revised cutoff was not associated with a statistical improvement in Youden’s J, and only minimally increased sensitivity. The impact of adjusting seroprevalence estimates in vaccinated NBDC donors for sensitivity and specificity was modest (Supplementary Table 3), not exceeding a proportional impact of 4% on point estimates. This indicates that the standard cutoff of the Ortho anti-N assay for VI detection performed sufficiently.

Recent receipt of primary or booster vaccinations reduced the likelihood of anti-N seroconversion following infection. Multivariable regression showed that recent receipt of an additional vaccine (booster) dose was associated with reduced detection, but the timing of primary vaccination relative to infection affected detection less. A Japanese study showed similar results, with reduced sensitivity to detect infection within 1-2 months of a 3^rd^ mRNA COVID-19 vaccine dose (sensitivity=78%), but high sensitivity for infections occurring >3 months post 2^nd^ or >4 months post 3^rd^ mRNA vaccine dose (25). The Moderna vaccine trial data (10), selected for VI occurring soon after vaccination and were collected relatively soon after infection, likely contributing to poor sensitivity in that study (10). Our data confirmed relatively poor sensitivity in specimens collected within one month of infection.

Since much of the lack of detection observed in this study occurred at times soon after infection, it is likely that in a longitudinal cohort many of the infections missed at the first post-infection specimen would be detected at later specimens from those individuals. Despite some waning of anti-N Ab levels following infection, longer-term durability of Ab detection over one year post-infection confirmed earlier findings by our group of robust durability of detection using anti-N total Ig Ag sandwich assays (15). Substantially reduced detection was observed in the first month following infection, especially during the first 14 days, with less than 50% of recently infected persons demonstrating anti-N seroconversion, but very good detection at later timepoints. Anti-N IgG assays used in numerous serosurveillance studies (16, 26–29) show more rapid waning in antibody signal than anti-N total Ig assays (15), and thus requires adjustments for seroreversion in estimating cumulative incidence (16). Although rapidly waning IgG assays may be less appropriate for serosurveillance aimed at cumulative incidence than total Ig assays, they may have advantages for detecting reinfections based on Ab boosting and as correlates of protection (30).

An important limitation of this study was that the case definition of infection in the validation data was based on self-reported diagnosed infections, without active surveillance of the cohort for asymptomatic infection. As a result, a large majority of survey-reported SCIs in the validation set were associated with COVID-19 symptoms (92%). In contrast, a meta-analysis of Omicron infections estimated 32.4% of infections were asymptomatic (31). This may result in a slight upward bias in overall sensitivity estimates. However, we found that adjusting for sensitivity to detect symptomatic and asymptomatic VI had a modest impact on seroprevalence estimates. A further limitation is that blood donors are not fully representative of the general population, including being generally healthier and more likely to be vaccinated and receive additional doses. Finally, vaccination and infection history were self-reported and not confirmed by healthcare records.

This study demonstrated that detection of first SARS-CoV-2 infections using the Ortho anti-N total Ig assay was robust in both vaccinated and unvaccinated donors, with overall sensitivities above 95%. It also found good durability of anti-N Ab detection for up to more than a year after infection. Seroprevalence studies using this assay can accurately estimate the proportion of people who have been infected with SARS-CoV-2 one or more times. Several factors impact the likelihood of anti-N seroconversion following first infection, including receipt of primary and additional vaccinations, sampling shortly after infection, and asymptomatic infection, although the impact of these factors was relatively small. Revising the cutoff improved sensitivity modestly use of the manufacturer recommended cutoff is likely appropriate for most serosurveillance studies.

## Funding

This work was supported by research contracts from the Centers for Disease Control and Prevention (CDC Contract 75D30120C08170). The findings and conclusions in this article are those of the authors and do not necessarily represent the views of the U.S. Centers for Disease Control and Prevention.

## Institutional Review Board Statement

All blood donors consented to use of deidentified, residual specimens for further research purposes. Consistent with the policies and guidance of the University of California–San Francisco Institutional Review Board, Vitalant Research Institute self-certified the use of deidentified donations in this study as not meeting the criteria for human subjects research. Centers for Disease Control and Prevention (CDC) investigators reviewed and relied on this determination as consistent with applicable federal law and CDC policy (45 C.F.R. part 46, 21 C.F.R. part 56; 42 U.S.C. § 241[d]; 5 U.S.C. § 552a; 44 U.S.C. § 3501). The donor surveys conducted by Vitalant and American Red Cross were conducted under protocols supervised and approved by the Advarra and American Red Cross Institutional Review Boards, respectively, and linked to biospecimens in deidentified form.

## Data Availability

Data used in the study may be shared, subject to IRB requirements and consent of all partnering organizations, upon reasonable request to the authors.

## Conflicts of Interest

Vitalant Research Institute receives research funding from QuidelOrtho. The authors have no other conflicts of interest to declare.

## Acknowledgements

The authors gratefully acknowledge CDC reviewers, whose comments greatly improved the manuscript. The contributions of numerous laboratory and data management staff, including Hasan Sulaeman, Brendan Balasko, Jahnavi Bhaskar, Patricia Villaflor, Kaya Duncan, Zhanna Kaidarova and Anh (Paul) Nguyen at Vitalant Research Institute, Marjorie D. Bravo at Vitalant, Edward P. Notari, James Haynes, Jamel Groves, and Gary Holley at American Red Cross, Rebecca Fink at Westat, Athena Nguyen, Dave Kovach, Chloe Byrne, Daishia Hall, and Tatum Fenner at Creative Testing Solutions and Melissa Briggs-Hagen at CDC, are also acknowledged with gratitude.

## Supplementary Materials

### Supplementary Appendix: Derivation of a revised non-reactive/reactive cutoff for the Ortho anti-nucleocapsid (anti-N) total Ig assay

To identify an optimal cutoff for serological detection of infection using first the Roche anti-N total Ig assay and later the Ortho anti-N total Ig assay, we performed receiver operating characteristic (ROC) curve analyses using samples from blood donors classified as not previously infected with SARS-CoV-2 (controls, ‘true negatives’) and previously infected with SARS-CoV-2 (cases, ‘true positives’). This analysis was performed based on samples collected cross-sectionally in the National Blood Donor Serosurvey (NBDS), without any clinical data or self-reported infection status, as described below.

To determine the optimal cutoff for the Roche anti-N total Ig assay, collected samples that had been tested in parallel with the Ortho anti-S and the Roche anti-N total Ig assays during monthly cross-sectional serosurveillance in the universal screening phase of the NBDS program (June to November 2020) were identified for optimization of the reactive/non-reactive cutoff of the Roche assay. Since samples were collected before widespread availability of vaccines, anti-S total Ig results were treated as independent indication of infection status (both positive and negative). The anti-S total Ig assay was treated as a ‘gold standard’ in this analysis, based on excellent performance demonstrated in a previous study (sensitivity of 95.8% and specificity of 100%), which was marginally better than the performance of the anti-N total Ig assay (1). Furthermore, the purpose of the analysis was to maximize sensitivity of the anti-N total Ig assay in the context of VI, which had not been assessed in the prior analysis. A total of 25,065 cases and 30,110 controls were identified using anti-S total Ig results (Roche optimization sample set). The controls were supplemented with 432 samples collected during 2019 before the emergence of SARS-CoV-2. A ROC curve analysis was performed, with optimality defined as maximal Youden’s J, i.e., an equal weighting of sensitivity and specificity. The optimal cutoff index (COI) on the Roche anti-N total Ig assay was COI≥0.205.

To determine the optimal cutoff for the Ortho anti-N total Ig assay, a similar sample set tested with the Ortho anti-N total Ig assay from before vaccine roll out was not available. NBDS samples collected after vaccines became available that had been tested with the Ortho anti-S total Ig assay and with both the Roche and Ortho anti-N total Ig assays were identified. For the ‘Ortho optimization sample set’ we identified 371 donors previously infected with SARS-CoV-2 (cases, i.e., ‘true positives’) based on reactivity on both anti-S total Ig (manufacturer’s cutoff) and Roche anti-N total Ig (revised cutoff derived above to improve sensitivity). For this analysis, controls (‘true negatives’) were restricted to pre-pandemic samples – 1,248 results supplied by QuidelOrtho from in-house specificity testing performed in support of the assay’s Emergency Use Authorization, supplemented with 200 pre-pandemic samples tested as part of a SARS-CoV-2 serology performance study (1). As in the analysis for the Roche assay, Youden’s J was used to identify an optimal cutoff. The optimal cutoff on the Ortho anti-N total Ig assay was S/CO≥0.395.

**Supplementary Figure 1** shows the distribution of S/CO values in the pre-pandemic control samples and the serologically defined cases (Ortho anti-S total Ig S/CO≥1 and Roche anti-N total Ig COI≥0.205), with vertical lines indicating the optimized cutoff based on our ROC analysis and the manufacturer’s recommended cutoff.

**Supplementary Figure 1.**
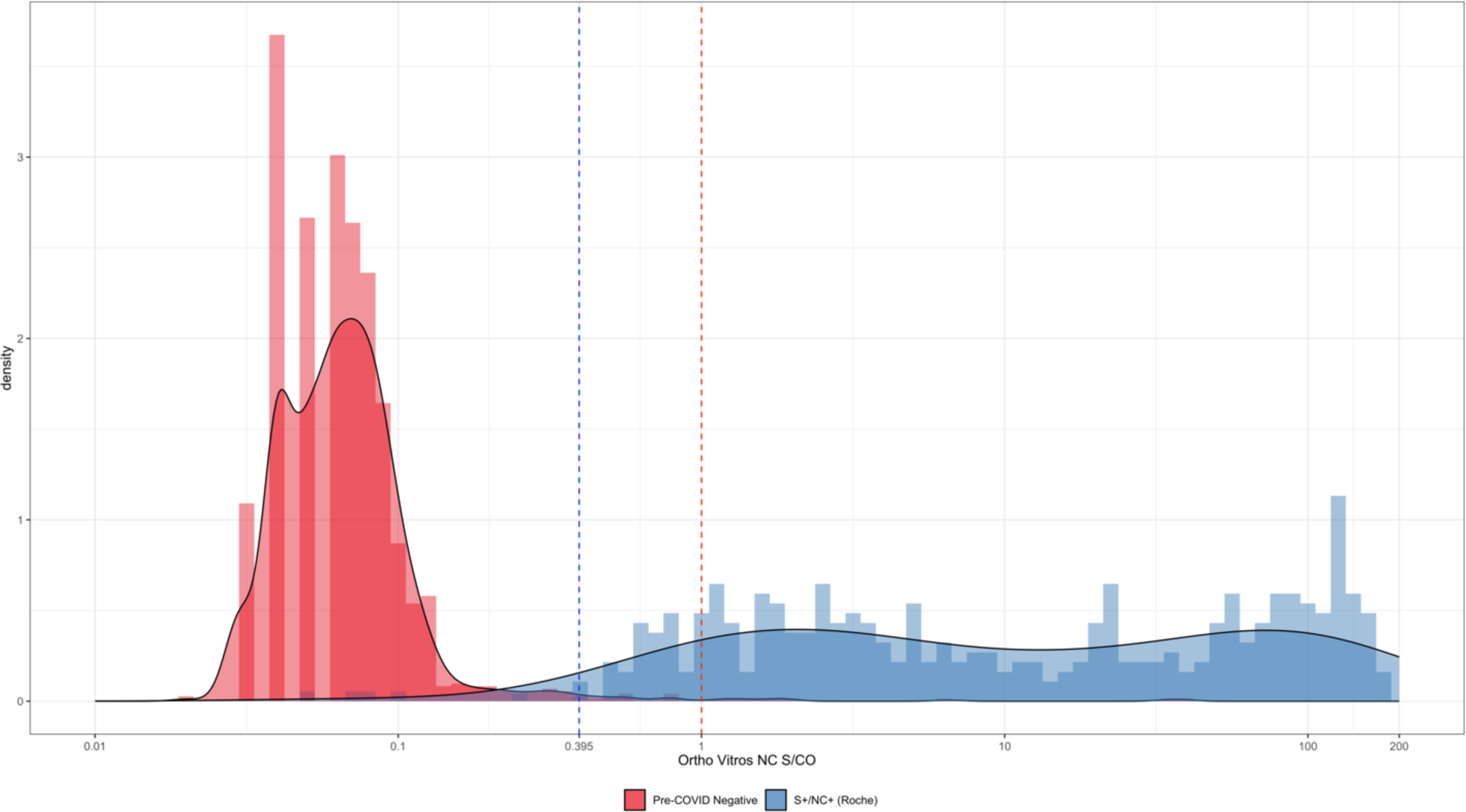
Anti-nucleocapsid (anti-N) signal-to-cutoff ratios on the Ortho anti-N total Ig assay, amongst pre-COVID-19 negative control samples, and samples from previously infected donors identified as anti-S and anti-N reactive using the Ortho anti-S and Roche anti-N total Ig assays. The red dashed line shows the manufacturer’s recommended cutoff and the blue dashed line shows revised cutoff identified as optimal using receiver operating characteristic (ROC) curve analysis.

**Supplementary Table 1.** Sensitivity of Ortho anti-nucleocapsid (anti-N) assay for detection of first infections in vaccinated donors. Proportion reactive in the first sample collected after reported swab-confirmed infection, collected 14 to 180 days post-infection, stratified by whether an anti-S reactive/anti-N nonreactive sample was observed after vaccination but before infection.

|  | First infections in vaccinated donors<br>(S+/N- observed following vaccination) |  |  | First infections in vaccinated donors<br>(No S+/N- following vaccination) |  |  |
| --- | --- | --- | --- | --- | --- | --- |
|  | N | Manufacturer's<br>cutoff*<br>% (95% CI) | Revised<br>cutoff**<br>% (95% CI) | N | Manufacturer's<br>cutoff*<br>% (95% CI) | Revised<br>cutoff**<br>% (95% CI) |
| Overall | 5079 | 95.5 (95.4, 95.6) | 97.1 (97.1, 97.2) | 3108 | 95.8 (95.7, 96.0) | 96.8 (96.7, 96.9) |
| Delta (Jul-Dec 2021) | 103 | 77.7 (56.0, 99.4) | 82.5 (65.6, 99.5) | 1246 | 95.3 (94.9, 95.6) | 96.3 (96.0, 96.6) |
| Omicron (Jan-Sep 2022) | 4976 | 95.9 (95.8, 96.0) | 97.4 (97.4, 97.5) | 1862 | 96.2 (96.0, 96.4) | 97.1 (96.9, 97.3) |
| Under 65 years | 3004 | 96.3 (96.2, 96.4) | 97.6 (97.6, 97.7) | 2190 | 96.0 (95.9, 96.2) | 96.8 (96.6, 96.9) |
| 65+ years | 2075 | 94.4 (94.1, 94.7) | 96.3 (96.2, 96.5) | 918 | 95.3 (94.8, 95.8) | 96.8 (96.5, 97.2) |
| Symptomatic*** | 4691 | 96.0 (95.9, 96.1) | 97.6 (97.6, 97.7) | 2725 | 96.5 (96.4, 96.6) | 97.4 (97.3, 97.5) |
| Asymptomatic*** | 322 | 89.4 (86.2, 92.7) | 90.4 (87.4, 93.4) | 305 | 90.8 (87.8, 93.8) | 92.1 (89.6, 94.7) |

\* S/CO $\geq 1.000$
\*\* S/CO $\geq 0.395$
\*\*\* Symptomatic/asymptomatic status could not be ascertained for all infections owing to incomplete survey responses

**Supplementary Table 2.** Factors influencing anti-nucleocapsid seroconversion following first swab-confirmed SARS-CoV-2 infection. The table shows the proportion of first post-infection samples that were reactive using the standard cutoff on the Ortho assay, the unadjusted odds ratio, and the adjusted odds ratio obtained from multivariable logistic regression.

| Variable | N | Percent detected | Unadjusted OR (95% CI) | Adjusted OR (95% CI) |
| --- | --- | --- | --- | --- |
| Overall | 14,094 | 93.9 |  |  |
| **Variant era** |  |  |  |  |
| Delta | 4,048 | 95.4 | 1.51 (1.28,1.78) | 0.80 (0.66,0.98) |
| Omicron | 10,046 | 93.3 | *ref* | *ref* |
| **Age Group (in years)** |  |  |  |  |
| 16-29 | 554 | 96.0 | 2.13 (1.37,3.31) | 1.83 (1.14,2.93) |
| 30-49 | 3,582 | 95.3 | 1.80 (1.49,2.18) | 1.41 (1.14,1.74) |
| 50-64 | 5,671 | 94.3 | 1.44 (1.24,1.69) | 1.23 (1.04,1.46) |
| 65+ | 4,287 | 91.9 | *ref* | *ref* |
| **Gender** |  |  |  |  |
| Female | 7,889 | 94.4 | 1.21 (1.05,1.38) | 1.04 (0.89,1.20) |
| Male | 6,205 | 93.3 | *ref* | *ref* |
| **Symptomatic infection** |  |  |  |  |
| Symptomatic | 12,613 | 94.4 | *ref* | *ref* |
| Asymptomatic | 1,260 | 88.9 | 0.47 (0.39,0.57) | 0.46 (0.37,0.57) |
| **Vaccination status\*** |  |  |  |  |
| Unvaccinated | 3,927 | 96.6 | *ref* |  |
| Partially vaccinated | 609 | 94.9 | 0.65 (0.43,0.97) |  |
| Fully vaccinated | 2,705 | 94.1 | 0.55 (0.43,0.70) |  |
| Boosted | 6,853 | 92.2 | 0.41 (0.34,0.50) |  |
| **Vaccination timing\*** |  |  |  |  |
| ≤1 months | 797 | 88.8 | 0.76 (0.58,1.00) |  |
| 2-3 months | 2,022 | 91.3 | *ref* |  |
| 4-6 months | 2,617 | 92.7 | 1.21 (0.98,1.50) |  |
| 7-9 months | 2,652 | 94.5 | 1.64 (1.30,2.05) |  |
| 10+ months | 2,079 | 94.0 | 1.50 (1.18,1.90) |  |
| **Vaccination status/timing (in days)** |  |  |  |  |
| Unvaccinated | 3,927 | 96.6 | *ref* | *ref* |
| Vaccinated (primary) ≤ 30 | 140 | 94.3 | 0.57 (0.28,1.20) | 0.57 (0.26,1.24) |
| Vaccinated (primary) 31-180 | 999 | 94.4 | 0.59 (0.43,0.81) | 0.63 (0.45,0.89) |
| Vaccinated (primary) > 180 | 2,175 | 94.1 | 0.56 (0.43,0.71) | 0.58 (0.44,0.75) |
| Vaccinated (boosted) ≤ 30 | 657 | 87.7 | 0.25 (0.19,0.33) | 0.29 (0.21,0.40) |
| Vaccinated (boosted) 31-180 | 3,640 | 91.4 | 0.37 (0.30,0.46) | 0.41 (0.32,0.52) |
| Vaccinated (boosted) > 180 | 2,556 | 94.4 | 0.58 (0.46,0.74) | 0.65 (0.50,0.85) |
| **Time to sample** |  |  |  |  |
| 0-13 days | 378 | 48.9 | 0.05 (0.04,0.06) | 0.04 (0.03,0.05) |
| 14-30 days | 2,109 | 87.9 | 0.34 (0.29,0.40) | 0.34 (0.28,0.40) |
| 31-90 days | 6,769 | 95.5 | *ref* | *ref* |
| 3-6 months | 3,539 | 97.5 | 1.85 (1.46,2.36) | 1.79 (1.40,2.28) |
| 7+ months | 1,299 | 98.1 | 2.38 (1.58,3.59) | 2.19 (1.44,3.35) |

\* Variables for vaccination status at time of infection, and for time from most recent vaccination to infection were replaced by a combined vaccination status and timing variable in multivariable logistic regression.

**Supplementary Table 3.** Impact of adjustment for sensitivity and specificity, for detection of VI, on estimated percentage of vaccinated donors who became infected during three time periods in the National Blood Donor Cohort. We assumed that 32.4% of VI were asymptomatic (2) and we adjusted for symptom status-specific sensitivity estimates (see Methods).

| **Time period** | **N infected / N tested\***<br>***among vaccinated,***<br>***not previously***<br>***infected*** | **Percentage infected**<br>**% (95% CI)**<br>***Naïve estimate*** | **Percentage infected**<br>**% (95% CI)**<br>***Adjusted for Se and***<br>***Sp*** | **Percentage infected**<br>**% (95% CI)**<br>***Adjusted for Se and***<br>***Sp, assuming 32.4%***<br>***of VI asymptomatic*** | **Difference between adjusted**<br>**and naïve estimate**<br>***Percentage points***<br>***(proportional difference)***<br>***Assuming 32.4% of VI***<br>***asymptomatic*** |
| --- | --- | --- | --- | --- | --- |
| From Q2 2021 to Q1 2022 | 7,656 / 33,815 | 22.64 (22.20–23.09) | 23.18 (22.60–23.76) | 23.52 (22.91–24.15) | 0.88 (3.9%) |
| From Q1 to Q2 2022 | 3,405 / 27,526 | 12.37 (11.99–12.76) | 12.37 (11.80–12.91) | 12.55 (11.96–13.11) | 0.18 (1.5%) |
| From Q2 to Q3 2022 | 5,569 / 24,147 | 23.06 (22.54–23.60) | 23.63 (22.96–24.28) | 23.98 (23.28–24.67) | 0.92 (4.0%) |

\* The numerator is the number of donors who were classified as vaccinated and not previously infected in the previous quarter, who were tested and seroconverted anti-N by the following quarter, and the denominator is the number that seroconverted plus the number who did not seroconvert, without any demographic weighting (3).

## Notes

### Author Declarations

All blood donors consented to use of deidentified, residual specimens for further research purposes. Consistent with the policies and guidance of the University of California San Francisco Institutional Review Board, Vitalant Research Institute self-certified the use of deidentified donations in this study as not meeting the criteria for human subjects research. Centers for Disease Control and Prevention (CDC) investigators reviewed and relied on this determination as consistent with applicable federal law and CDC policy (45 C.F.R. part 46, 21 C.F.R. part 56; 42 U.S.C. 241[d]; 5 U.S.C. 552a; 44 U.S.C. 3501). The donor surveys conducted by Vitalant and American Red Cross were conducted under protocols supervised and approved by the Advarra and American Red Cross Institutional Review Boards, respectively, and linked to biospecimens in deidentified form.

## References

1. Jones JM, Manrique IM, Stone MS, Grebe E, Saa P, Di Germanio C, et al. Estimates of SARS-CoV-2 Seroprevalence and Incidence of Primary SARS-CoV-2 Infections Among Blood Donors, by COVID-19 Vaccination Status - United States, April 2021-September 2022. MMWR Morbidity and mortality weekly report. 2023;72(22):601–5.

2. Busch MP, Stone M. Serosurveillance for Severe Acute Respiratory Syndrome Coronavirus 2 (SARS-CoV-2) Incidence Using Global Blood Donor Populations. Clinical infectious diseases : an official publication of the Infectious Diseases Society of America. 2021;72(2):254–6.

3. O’Brien SF, Lieshout-Krikke RW, Lewin A, Erikstrup C, Steele WR, Uzicanin S, et al. Research initiatives of blood services worldwide in response to the covid-19 pandemic. Vox Sang. 2021;116(3):296–304.

4. Stone M. SARS-CoV-2 infection incidence and vaccination among repeat blood donors: Protocol for a multi-site prospective cohort in the US (2021-2022). Forthcoming. 2023.

5. Stone M, Di Germanio C, Wright DJ, Sulaeman H, Dave H, Fink RV, et al. Use of US Blood Donors for National Serosurveillance of Severe Acute Respiratory Syndrome Coronavirus 2 Antibodies: Basis for an Expanded National Donor Serosurveillance Program. Clinical infectious diseases : an official publication of the Infectious Diseases Society of America. 2022;74(5):871–81.

6. Jones JM, Stone M, Sulaeman H, Fink RV, Dave H, Levy ME, et al. Estimated US Infection- and Vaccine-Induced SARS-CoV-2 Seroprevalence Based on Blood Donations, July 2020-May 2021. Jama. 2021;326(14):1400–9.

7. Jones JM, Opsomer JD, Stone M, Benoit T, Ferg RA, Stramer SL, et al. Updated US Infection- and Vaccine-Induced SARS-CoV-2 Seroprevalence Estimates Based on Blood Donations, July 2020-December 2021. Jama. 2022;328(3):298–301.

8. U.S. Centers for Disease Control and Prevention. 2022 Nationwide COVID-19 Infection- and Vaccination-Induced Antibody Seroprevalence (Blood donations) 2023 [Available from: https://covid.cdc.gov/covid-data-tracker/#nationwide-blood-donor-seroprevalence-2022.

9. Whitaker HJ, Gower C, Otter AD, Simmons R, Kirsebom F, Letley L, et al. Nucleocapsid antibody positivity as a marker of past SARS-CoV-2 infection in population serosurveillance studies: impact of variant, vaccination, and choice of assay cut-off. medRxiv. 2021:2021.10.25.21264964.

10. Follmann D, Janes HE, Buhule OD, Zhou H, Girard B, Marks K, et al. Antinucleocapsid Antibodies After SARS-CoV-2 Infection in the Blinded Phase of the Randomized, Placebo-Controlled mRNA-1273 COVID-19 Vaccine Efficacy Clinical Trial. Ann Intern Med. 2022;175(9):1258–65.

11. Dhakal S, Yu T, Yin A, Pisanic N, Demko ZO, Antar AAR, et al. Reconsideration of anti-nucleocapsid IgG antibody as a marker of SARS-CoV-2 infection post-vaccination for mild COVID-19 patients. Open Forum Infectious Diseases. 2022.

12. Dalai SC, Dines JN, Snyder TM, Gittelman RM, Eerkes T, Vaney P, et al. Clinical Validation of a Novel T-Cell Receptor Sequencing Assay for Identification of Recent or Prior Severe Acute Respiratory Syndrome Coronavirus 2 Infection. Clinical infectious diseases : an official publication of the Infectious Diseases Society of America. 2022;75(12):2079–87.

13. Zuo J, Dowell AC, Pearce H, Verma K, Long HM, Begum J, et al. Robust SARS-CoV-2-specific T cell immunity is maintained at 6 months following primary infection. Nat Immunol. 2021;22(5):620–6.

14. Anderson M, Stec M, Gosha A, Mohammad T, Boler M, Tojo Suarez R, et al. Longitudinal Severe Acute Respiratory Syndrome Coronavirus 2 Vaccine Antibody Responses and Identification of Vaccine Breakthrough Infections Among Healthcare Workers Using Nucleocapsid Immunoglobulin G. The Journal of infectious diseases. 2022;226(11):1934–42.

15. Stone M, Grebe E, Sulaeman H, Di Germanio C, Dave H, Kelly K, et al. Evaluation of Commercially Available High-Throughput SARS-CoV-2 Serologic Assays for Serosurveillance and Related Applications. Emerg Infect Dis. 2022;28(3):672–83.

16. Buss LF, Prete CA, Jr., Abrahim CMM, Mendrone A, Jr., Salomon T, de Almeida-Neto C, et al. Three-quarters attack rate of SARS-CoV-2 in the Brazilian Amazon during a largely unmitigated epidemic. Science. 2021;371(6526):288–92.

17. Erikstrup C, Laksafoss AD, Gladov J, Kaspersen KA, Mikkelsen S, Hindhede L, et al. Seroprevalence and infection fatality rate of the SARS-CoV-2 Omicron variant in Denmark: A nationwide serosurveillance study. Lancet Reg Health Eur. 2022;21:100479.

18. Honge BL, Hindhede L, Kaspersen KA, Harritshoj LH, Mikkelsen S, Holm DK, et al. Long-term detection of SARS-CoV-2 antibodies after infection and risk of re-infection. Int J Infect Dis. 2022;116:289–92.

19. Sulaeman H, Grebe E, Dave H, McCann L, Di Germanio C, Sanghavi A, et al. Evaluation of Ortho VITROS and Roche Elecsys S and NC Immunoassays for SARS-CoV-2 Serosurveillance Applications. Microbiol Spectr. 2023:e0323422.

20. Fink RV, Fisher L, Sulaeman H, Dave H, Levy ME, McCann L, et al. How do we…form and coordinate a national serosurvey of SARS-CoV-2 within the blood collection industry? Transfusion. 2022;62(7):1321–33.

21. Busch MP, Stramer SL, Stone M, Yu EA, Grebe E, Notari E, et al. Population-Weighted Seroprevalence From Severe Acute Respiratory Syndrome Coronavirus 2 (SARS-CoV-2) Infection, Vaccination, and Hybrid Immunity Among US Blood Donations From January to December 2021. Clinical infectious diseases : an official publication of the Infectious Diseases Society of America. 2022;75(Suppl 2):S254–S63.

22. Lambrou AS, Shirk P, Steele MK, Paul P, Paden CR, Cadwell B, et al. Genomic Surveillance for SARS-CoV-2 Variants: Predominance of the Delta (B.1.617.2) and Omicron (B.1.1.529) Variants - United States, June 2021-January 2022. MMWR Morbidity and mortality weekly report. 2022;71(6):206–11.

23. Shang W, Kang L, Cao G, Wang Y, Gao P, Liu J, et al. Percentage of Asymptomatic Infections among SARS-CoV-2 Omicron Variant-Positive Individuals: A Systematic Review and Meta-Analysis. Vaccines (Basel). 2022;10(7).

24. Rogan W, Gladen E. Estimating the Prevalence from the Results of a Screening Test. American Journal of Epidemiology. 1978;107(1):71–6.

25. Mizoue T, Yamamoto S, Konishi M, Oshiro Y, Inamura N, Nemoto T, et al. Sensitivity of anti-SARS-CoV-2 nucleocapsid protein antibody for breakthrough infections during the epidemic of the Omicron variants. The Journal of infection. 2022;85(5):573–607.

26. Buss LF, Sabino EC. Intense SARS-CoV-2 transmission among affluent Manaus residents preceded the second wave of the epidemic in Brazil. Lancet Glob Health. 2021;9(11):e1475–e6.

27. Bloch EM, Kyeyune D, White JL, Ddungu H, Ashokkumar S, Habtehyimer F, et al. SARS-CoV-2 seroprevalence among blood donors in Uganda: 2019-2022. Transfusion. 2023;63(7):1354–65.

28. He Z, Ren L, Yang J, Guo L, Feng L, Ma C, et al. Seroprevalence and humoral immune durability of anti-SARS-CoV-2 antibodies in Wuhan, China: a longitudinal, population-level, cross-sectional study. Lancet. 2021;397(10279):1075-84.

29. Murhekar MV, Bhatnagar T, Thangaraj JWV, Saravanakumar V, Santhosh Kumar M, Selvaraju S, et al. Seroprevalence of IgG antibodies against SARS-CoV-2 among the general population and healthcare workers in India, June-July 2021: A population-based cross-sectional study. PLoS Med. 2021;18(12):e1003877.

30. Prete CA, Jr., Buss LF, Buccheri R, Abrahim CMM, Salomon T, Crispim MAE, et al. Reinfection by the SARS-CoV-2 Gamma variant in blood donors in Manaus, Brazil. BMC Infect Dis. 2022;22(1):127.

31. Von Bartheld CS, Wang L. An Explanation for Reports of Increased Prevalence of Olfactory Dysfunction with Omicron: Asymptomatic Infections. The Journal of infectious diseases. 2023.

## Supplementary References

1. Stone M, Grebe E, Sulaeman H, Di Germanio C, Dave H, Kelly K, et al. Evaluation of Commercially Available High-Throughput SARS-CoV-2 Serologic Assays for Serosurveillance and Related Applications. Emerg Infect Dis. 2022;28(3):672–83.

2. Shang W, Kang L, Cao G, Wang Y, Gao P, Liu J, et al. Percentage of Asymptomatic Infections among SARS-CoV-2 Omicron Variant-Positive Individuals: A Systematic Review and Meta-Analysis. Vaccines (Basel). 2022;10(7).

3. Jones JM, Manrique IM, Stone MS, Grebe E, Saa P, Di Germanio C, et al. Estimates of SARS-CoV-2 Seroprevalence and Incidence of Primary SARS-CoV-2 Infections Among Blood Donors, by COVID-19 Vaccination Status -United States, April 2021-September 2022. MMWR Morbidity and mortality weekly report. 2023;72(22):601–5.

